# Oncogenomic profiling of cutaneous pericytic tumours reveals distinct drivers and shared biological processes

**DOI:** 10.1101/2025.07.31.25332530

**Authors:** Martin Del Castillo Velasco-Herrera, Saamin Cheema, Kim Wong, Jamie Billington, Ian Vermes, Elizabeth Anderson, Elizabeth Ferla, Paul W. Harms, Nicolas de Saint Aubain, Emily L. Clarke, William Merchant, Ahmed K. Alomari, Neil Rajan, Peter Ferguson, Maximillian A. Weigelt, Carlos Monteagudo, Steven D. Billings, Mark J. Arends, Ingrid Ferreira, Thomas Brenn, Louise van der Weyden, David J. Adams

## Abstract

To-date the genomic landscape of cutaneous pericytic tumours (PTs), which represent a morphological continuum, have not been comprehensively explored. In order to identify the driver events of PTs from across their histological spectrum, and potentially aid the current classification system, we sequenced DNA (whole-exome) and RNA (pulldown transcriptome) from tumour-normal pairs classified by two different dermatopathologists of angioleiomyoma (n=37), glomus tumour (n=30) and myopericytoma (n=11); with all sequencing data deposited in the European Genome and Phenome Archive.

---

The tumour mutational burden of all three tumour types was low with the exception of a single glomus tumour (GT; PD56659a), in which COSMIC mutational signatures SBS7a/b were identified, suggesting UV exposure (Fig. 1a). Recurrently mutated genes in angioleiomyoma (ALM) included *PIK3CA* and *MAPK1* (2/37 each, with one tumour having three mutations in *PIK3CA*; Fig. 1a). *PIK3CA* was identified as a significantly mutated gene (using two independent algorithms; q-value<0.1), suggesting it is a driver gene of ALM. In agreement with a previous report,^1^ we found an ALM with a *PDGFRB* mutation. However, we did not observe the previously reported *NOTCH3* mutations,^1^ although we did observe mutations in other *NOTCH* family members. Recurrently mutated genes in GT included *NF1* and *PCLO* (3/30 each; PD56659a having two mutations in each gene), and *PDGFRB* and *TEK* (2/30 each; Fig. 1a). *NF1* was identified as a significantly mutated gene (using two independent algorithms; q-value<0.1), suggesting it is a driver gene of GT. In agreement with a previous report,^1^ we found GTs with mutations in *PDGFRB* or *NOTCH3*. Recurrently mutated genes in myopericytoma (MPC) included *LRP1B* (2/11, 18%; Fig. 1a). In agreement with previous reports,^1,2^ we found MPCs with mutations in *PDGFRB* or *NOTCH3*. One study reported the presence of *BRAF*^*V600E*^ in a subset of MPC, however, this has not been replicated in later studies,^2,3^ consistent with our findings.

**Fig. 1.**
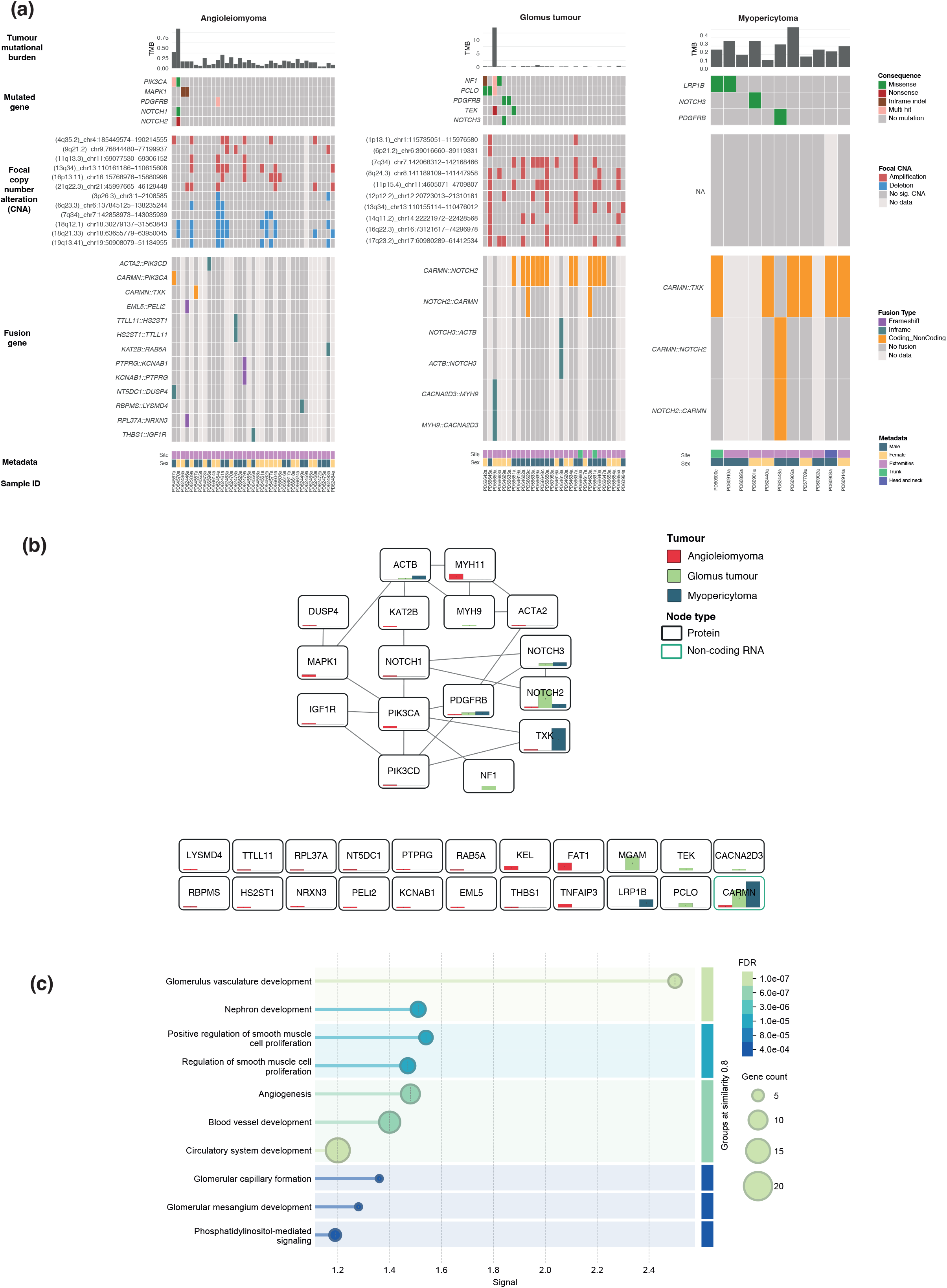
Overall mutational landscape of pericytic tumours. a) For each of the three tumour types, the panels show the tumour mutational burden, recurrently mutated genes (n≥2) or previously reported mutated genes/families, significant focal (<10Mb) copy number alterations (listing any known cancer-associated genes within the amplification or deletion), fusion genes, associated metadata (tumour site and patient sex) and sample ID. b) Protein-Protein interaction network identified using STRING v12.0 amongst all the altered genes in PTs. Each node contains bar plots with the proportion of samples analysed per tumour type that contain alterations, by either mutations, gene fusions or CNAs y-axis 0 to 70%. c) Gene ontology enrichment analysis showing the statistically significant biological processes like, angiogenesis, positive regulation of smooth muscle cells, and blood vessel development are associated to the genes found to be altered in PTs.

Considering copy number (CN) alterations as potential driver events of pericytic tumours (PT), we focussed on statistically significantly focal (<10Mb) gains (amplifications) and losses (deletions; Fig. 1). ALM showed regions of significant CN gain (n=6) and loss (n=6), some found in up to 25% of tumours, and four regions encompassing known cancer-associated genes; *FAT1* in the 4.7 Mb amplification at 4q35.2, *MYH11* in the 112 kb amplification at 16p13.11, *TNFAIP3* in the 390 kb deletion at 6q23.3, and *KEL* in the 177 kb deletion at 7q34. GT showed ten regions of significant CN gain, some found in up to 30% of tumours, with the 100 kb amplification at 7q34 encompassing the cancer-associated gene, *MGAM* (Fig. 1a). MPC did not show any significant CN alterations.

As fusion genes are driver events in many tumour types, we looked for their presence in PTs, particularly focussing on recurrent fusions. Thirteen fusions were identified in the ALM cohort, including *THBS1::IGF1R* which was recently identified as a novel fusion gene in a myopericytic tumour,^4^ however, none were recurrent (Fig. 1a). In contrast, the *CARMN::NOTCH2* fusion (previously known as *MIR143::NOTCH2*) was found in 13/18 (72%) of GT cases with RNAseq data (two cases also carrying the reciprocal fusion; Fig. 1a), consistent with previous reports of this fusion being found in both benign and malignant GT of cutaneous (extremities), visceral and soft tissue origin (36% - 76%; frequencies vary by site).^3,5^ However, it is important to note that whilst a previous study did not find the *CARMN::NOTCH2* fusion in any MPC (0/6) or ALM (0/18) samples,^5^ we found *CARMN::NOTCH2* (and its reciprocal fusion) in a single MPC sample, suggesting its presence is not exclusive to GT. It is worth noting that this particular case showed histological features of both MPC and GT. In the MPC cohort, the *CARMN::TKX* fusion was found in 6/7 (86%) cases, with one case showing a *CARMN::NOTCH2* fusion instead. However, is important to note that the *CARMN::TKX* fusion was also present in a single ALM sample (with confirmed histological presence of thick muscular bundles) which is consistent with previous reports,^6^ thus its presence is not exclusive to MPC.

In summary, our comprehensive characterisation of the genomic landscape of a large cohort of PTs has identified distinct driver events between the different tumour types, despite a morphological spectrum; the *CARMN::TXK* fusion gene in MPC, the *CARMN::NOTCH2* fusion gene, CN gains and *NF1* mutations in GT, and CN deletions/gains and *PIK3CA* mutations in ALM. Importantly, the *CARMN::TXK* and *CARMN::NOTCH2* fusion genes were not exclusive to a particular tumour subtype which is relevant when considering these events as diagnostic markers. However, when considering all the genes altered in the PTs (either by mutation, copy number alteration or fusion partner), there is some commonality in terms of shared protein interactions and biological processes (Fig. 1b-c), in agreement with their morphological overlap.

## Data Availability

All sequencing data deposited in the European Genome and Phenome Archive.

## Funding sources

This study was funded by the Medical Research Council (MR/V000292/1) and Wellcome Trust (220540/Z/20/A). NR is supported by the Newcastle NIHR Biomedical Research Centre. For the purpose of Open Access, the author has applied a CC BY public copyright licence to any Author Accepted Manuscript version arising from this submission.

## Conflicts of interest

The authors have no competing or financial interests to declare.

## Notes

### Competing Interest Statement

The authors have declared no competing interest.

### Author Declarations

Ethical approval for the use of all patient samples in this project was obtained by a committee at the institution of origin and from Research Governance at the Wellcome Sanger Institute. This study is part of the DERMATLAS Project that has been approved by the NHS Health Research Authority; Research Ethics Committee (REC) reference: 21/PR/1024, IRAS project ID: 304621.

